# Understanding diagnostic delay for endometriosis: a scoping review

**DOI:** 10.1101/2024.01.08.24300988

**Authors:** Jodie Fryer, Amanda J. Mason-Jones, Amie Woodward

## Abstract

**Introduction:** Diagnostic delay for endometriosis is a well-established phenomenon. Despite this, little is known about where in the health care system these delays occur or why they occur. Our review is the first attempt to synthesise and analyse this evidence.

**Methods:** A systematic scoping review with a pre-specified protocol was used to incorporate the global mixed methods literature on diagnostic delay for endometriosis. Four databases (PubMed, MEDLINE, EMBASE, PsychINFO) were searched from inception to September 2023 with a search strategy designed specifically for each.

**Results:** The search yielded 367 studies, 22 of which met the inclusion criteria. A third of studies has been published since 2020 and 65% were from high income countries. Six were qualitative and 16 were quantitative studies. The average age of onset of endometriosis was 14 years for adolescents and 20 for adults. On average, the diagnostic delay reported for endometriosis across the included studies was 6.6 years (range 1.5 to 11.3 years) but this masked the very wide differences reported between countries such as a 0.5-year delay in Brazil to a 27-year delay in the UK.

**Discussion:** Health system barriers included access to private healthcare for those with limited finance, physical access for those using public health systems and a general lack of knowledge amongst patients and health care professionals. Women often reported feeling unheard by health professionals. Considering the impact on individuals and the health system, addressing diagnostic delay for endometriosis must remain a priority for researchers, health care providers and policy makers.

**What is already known on this topic:** Endometriosis is currently difficult to diagnose. This results in delays in diagnosis which negatively impacts those suffering and increases the severity of pain and extent of the disease with increased costs to health systems.

**What this study adds:** The scoping review methodology included studies using a range of methods. The longest average delay occurs in secondary care. Those seeking public health care experienced longer average delay in diagnosis compared to those seeking private health care. Improved clinical guidelines may reduce diagnostic delay.

**How this study might affect research, practice or policy:** This is the first known review to explore diagnostic delay for endometriosis and provides an overview of the current literature. Clearer definitions of diagnostic delay for endometriosis are needed to aid in comparisons across countries. Improving education, tracking outcomes through medical records and developing non-invasive diagnostic tools will be crucial to improve women’s health.

## Introduction

Endometriosis is an oestrogen dependent gynaecological condition characterised by the presence of active endometrial tissue lying outside of the uterus, typically in the pelvic region (1). It is a chronic, progressive inflammatory disease which affects more than 170 million women worldwide (2). Endometriosis mainly affects women of reproductive age (15-49 years), with up to 1 in 10 believed to have the condition, although it is estimated that as many as 60% of endometriosis cases remain undiagnosed (2, 3). Prevalence estimates of endometriosis are generally poor and highly varied, ranging from 4% to 50%; however the most consistent estimates suggest prevalence ranging from 6-10% (4). Despite the progressive nature of endometriosis, a correct diagnosis takes an average of 10 years and at least 7 visits to a health practitioner (5, 6). This lengthy delay is reflected in the disease burden in which gynaecological diseases are reported as the leading cause of Disability Adjusted Life Years (DALYs) and Years Lived with Disability (YLD) among the 15-49-year age group (7). This is despite clear clinical diagnostic indicators including chronic pelvic pain (CPP), dysmenorrhea (painful, heavy menstruation), dyspareunia (painful intercourse), that are known for 82.9% of women (1, 8). Apart from the YLD the economic impact includes increased costs to the individual, to healthcare providers, and to the wider economic infrastructure (9). The current ‘gold standard’ for diagnosis is a laparoscopy, although surgeons may be hesitant to perform this due to the invasive nature of the procedure (8, 10). There is also evidence that symptoms may be dismissed as ‘normal’ by health care practitioners (1, 11).

The aim of this review was to explore the delay faced by those attempting to obtain a diagnosis of endometriosis and appropriate treatment.

## Methods

The study protocol was registered on the Open Science Framework OSF: 10.31219/osf.io/yzuvb

### Patient and public involvement

Women who have experienced diagnostic delay for endometriosis were involved in designing the research. The research question was informed by their priorities, experiences and preferences. Dissemination of this research will be facilitated through charities focussed on endometriosis.

### Data sources and search strategy

Development of the search strategy was guided by the SPIDER framework to ensure key concepts were captured in searches. Four databases were searched between from inception to September 2023. They included PubMed, MEDLINE, EMBASE and PsycINFO. No date limits were set on the searches. Search terms included key terms derived from search strings relating to ‘endometriosis’ and ‘diagnostic delay’ and were adapted for each database; For example, the search strategy for MEDLINE was: ‘Endometriosis.mp. or (exp Pelvic Pain/ or exp Chronic Pain/)) and exp Delayed Diagnosis/’.

### Eligibility criteria

Included studies were primary research in English involving the pelvic region or reproductive organs only, that mentioned pelvic pain with a suspicion of endometriosis, and diagnostic delay (in the context of endometriosis).

### Screening and data extraction

All studies were screened by one reviewer (JF) with a 10 percent sample checked by a second reviewer (MS) and any disagreements resolved by a third reviewer (AW/AMJ).

Data were extracted on a predeveloped and piloted data extraction form and included study characteristics, methods and design, and demographic characteristics of the population. Additionally, the most frequently reported symptoms, length of and reason for delay were recorded.

### Analysis

Studies were grouped by themes that emerged from the individual included studies (12) and contextualised to form a public policy perspective using the socio-ecological model (13).

## Results

### Selection of studies

The searches yielded 367 studies following deduplication. Title and abstract screening, and full-text screening resulted in 23 studies that met the inclusion criteria (see figure 1).

**Figure 1.**
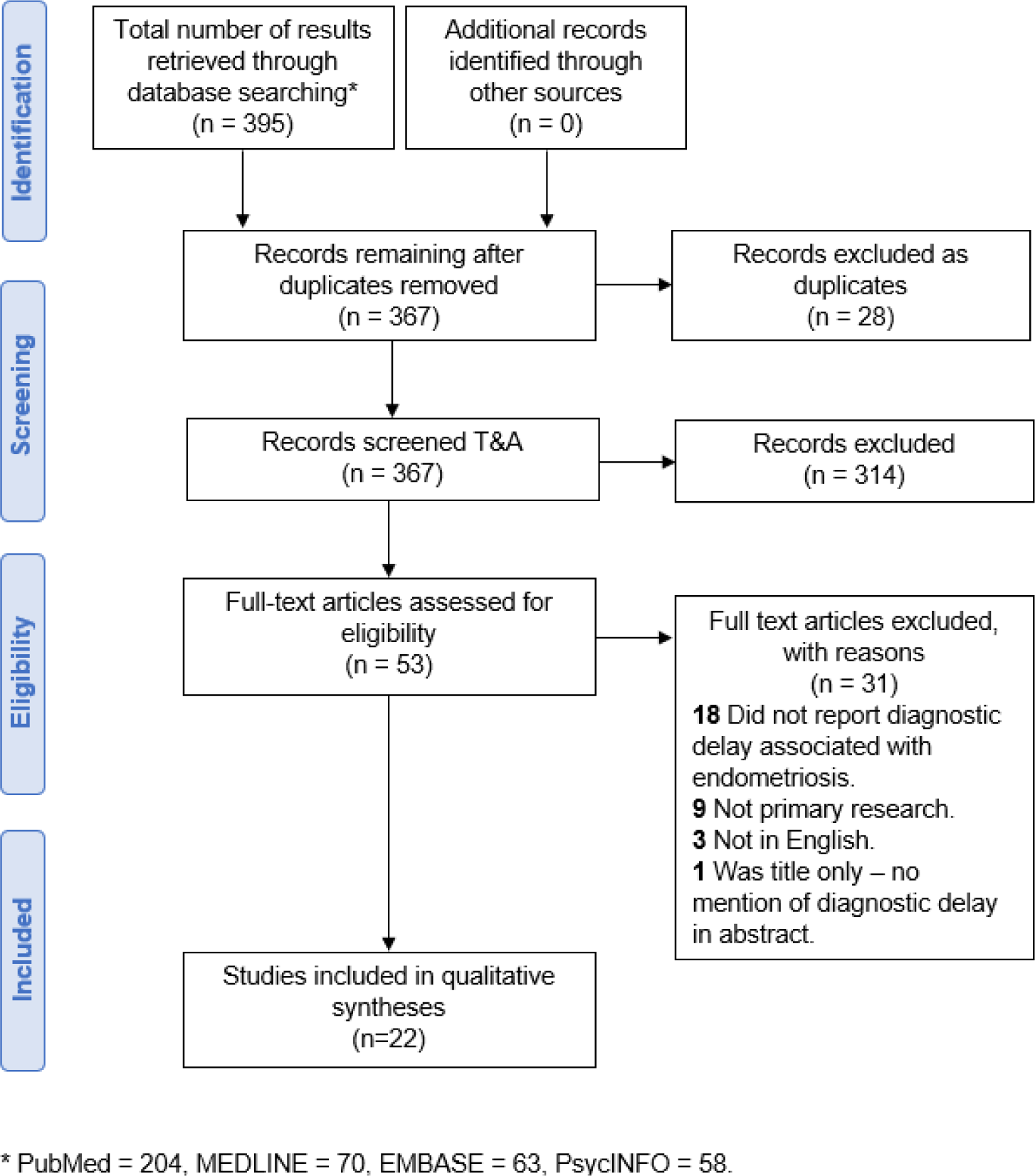
PRISMA flow diagram.

No formal quality appraisal was undertaken in line with methodological guidance for scoping reviews (14).

### Study characteristics

Table 1 provides an overview of the included studies and highlights the diversity of methods used. Six were qualitative and 16 were quantitative studies. Almost a third of studies (8/22) were published relatively recently (since 2020) from a range of countries. Fifteen were conducted in high income countries including the UK (15, 16), US (17–21), Netherlands (22, 23), Norway (24, 25), Canada (26), Australia (27), New Zealand (28), and Italy (29). Three were conducted in middle income countries; Brazil (30, 31) and Iran (32) and four were conducted in multiple countries (33–36). The average age of participants across the studies was 32.7 but the age range of participants was between 12 and 74 years old.

**Table 1:**
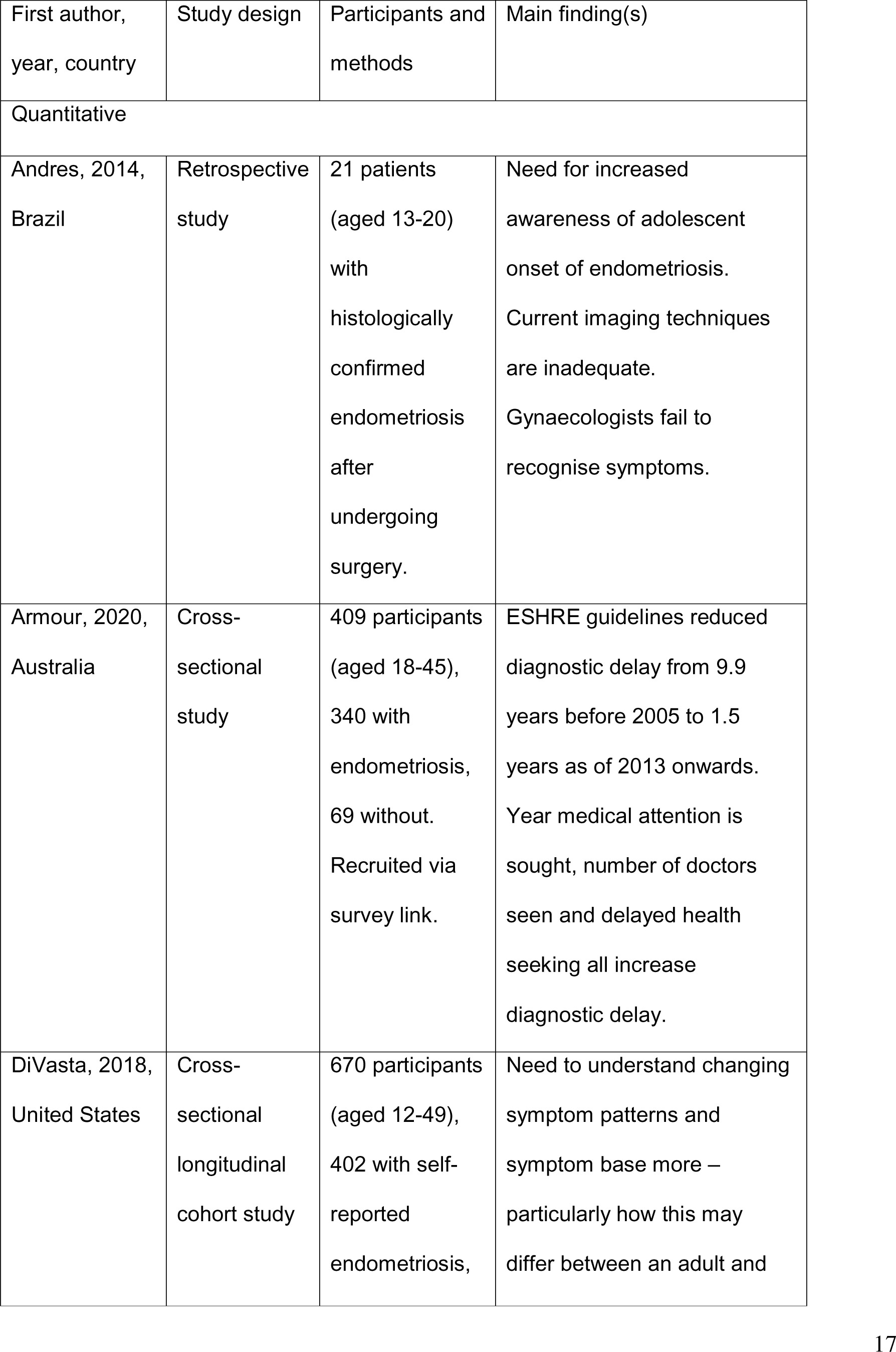
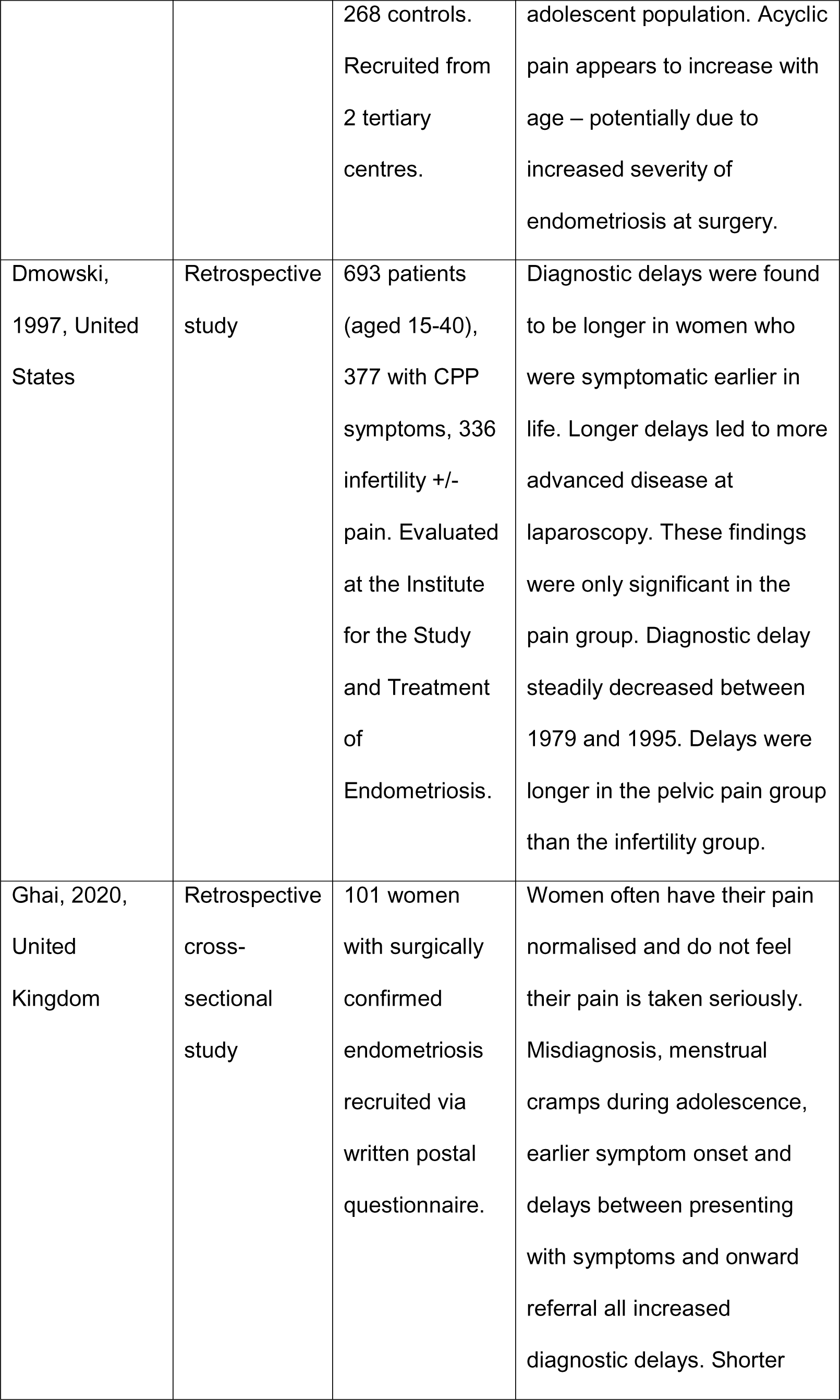
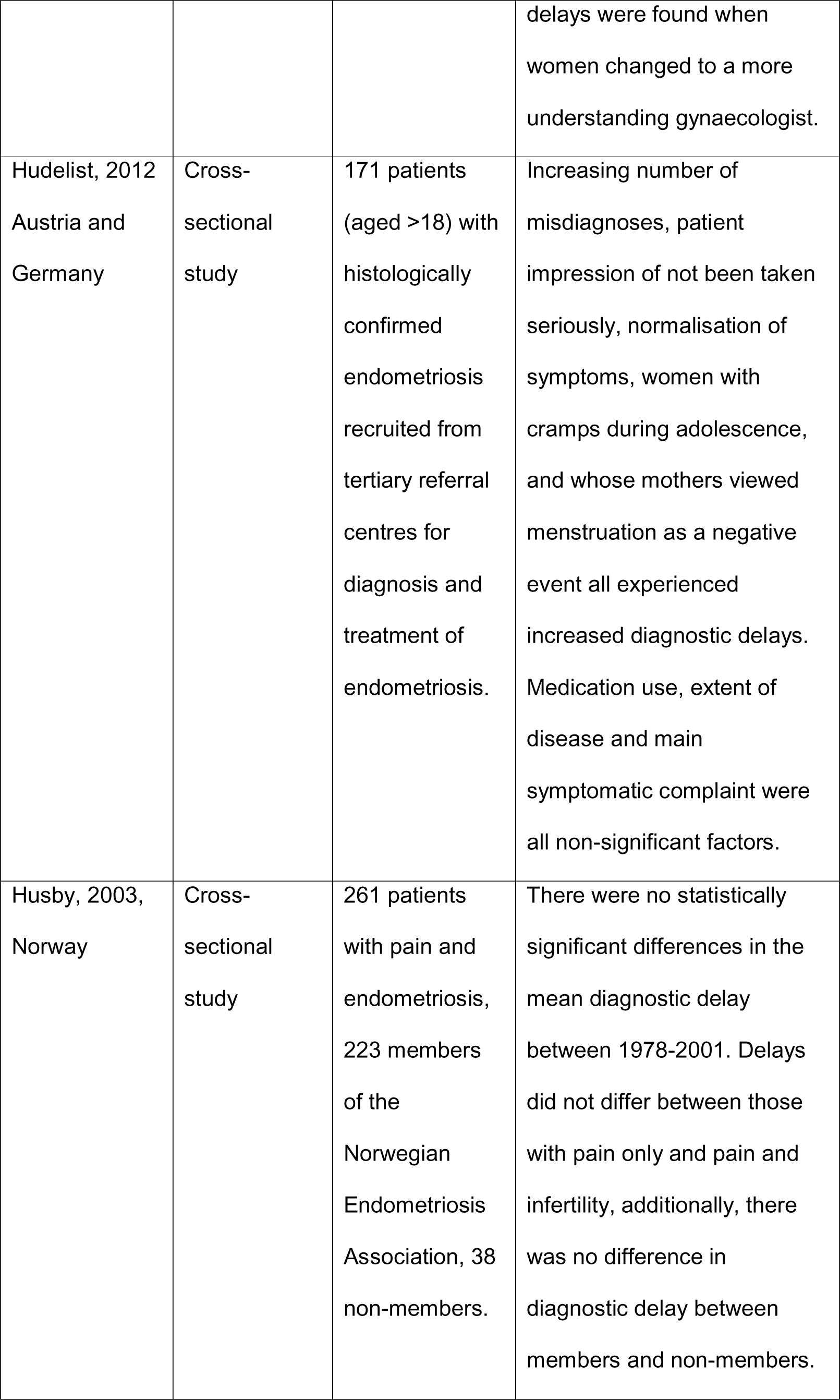
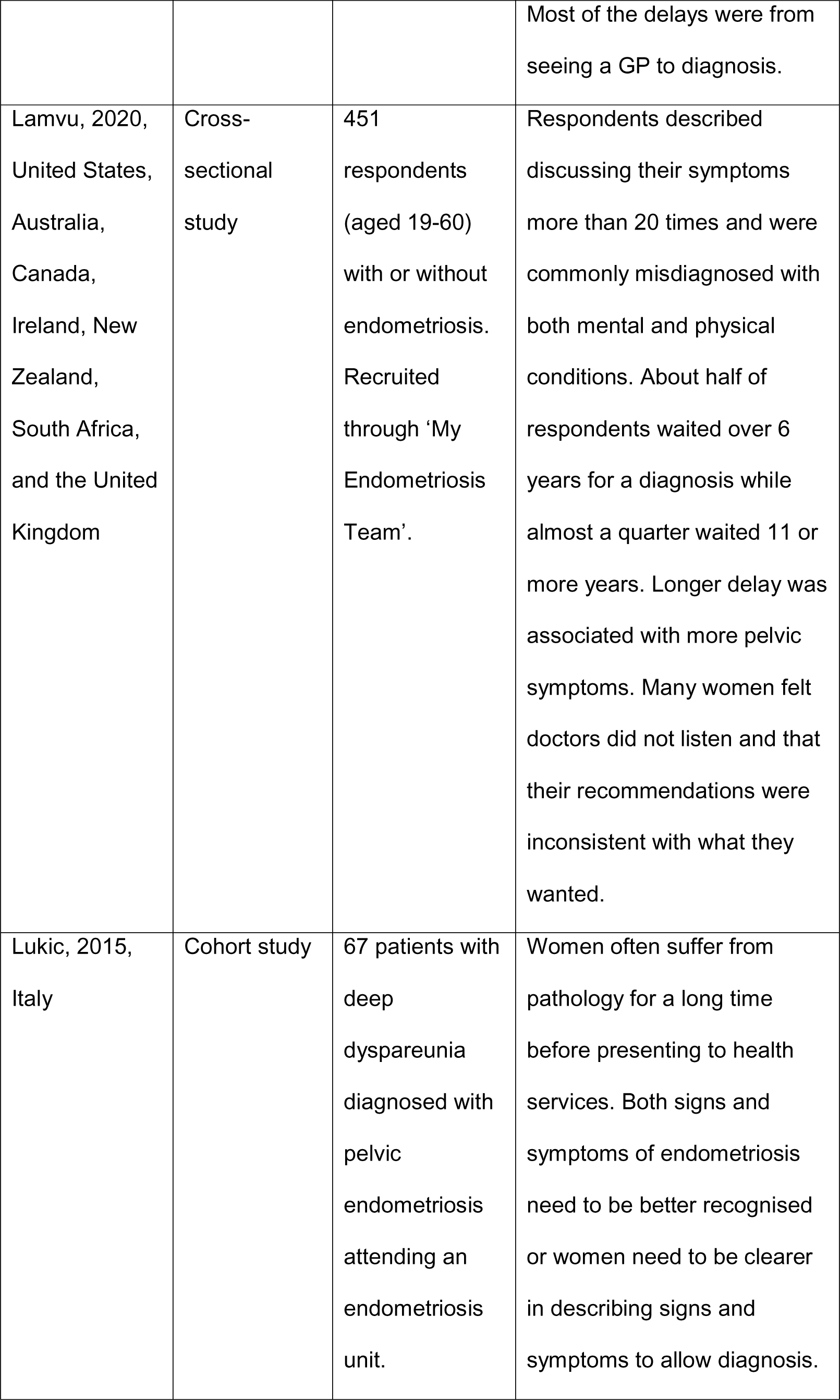
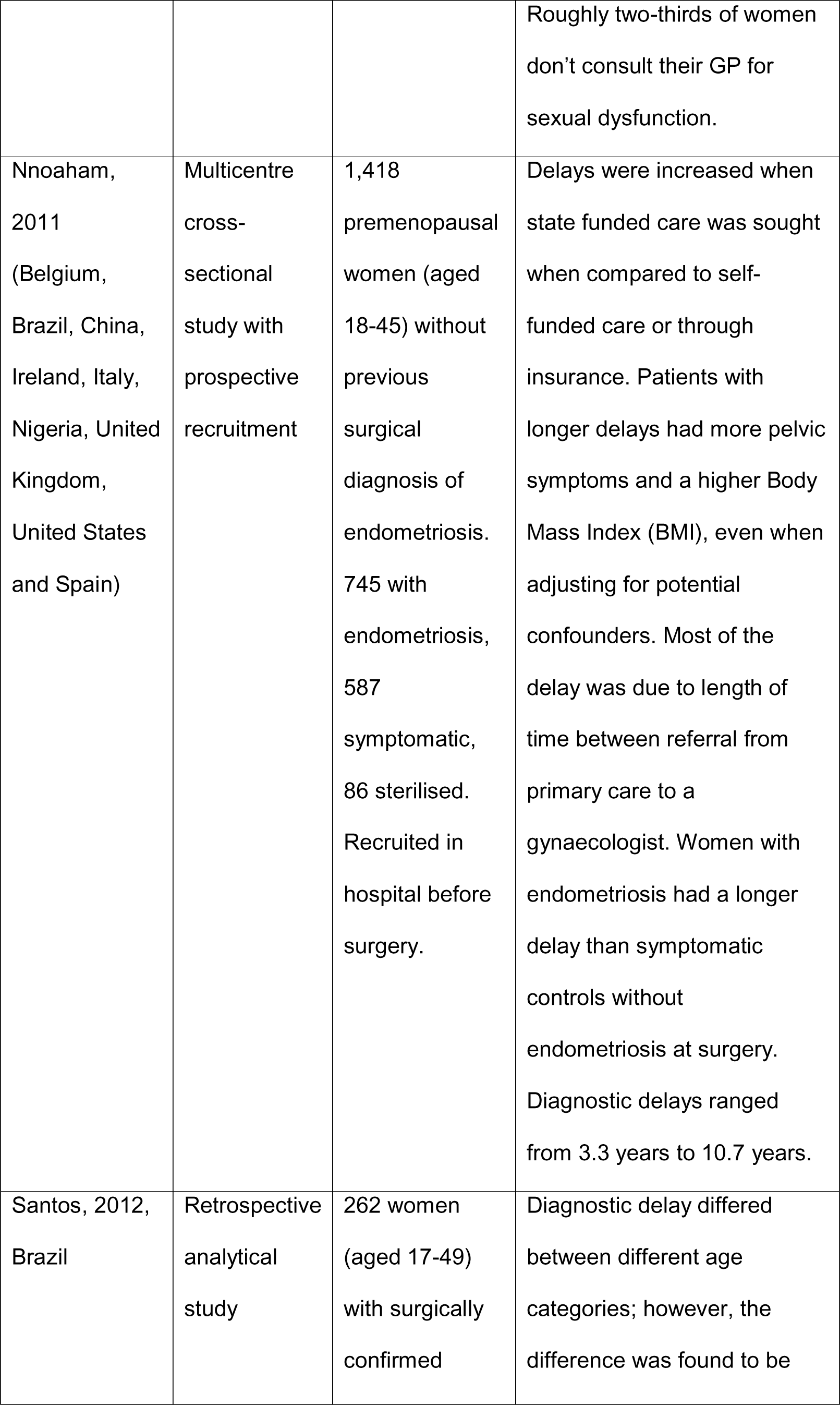
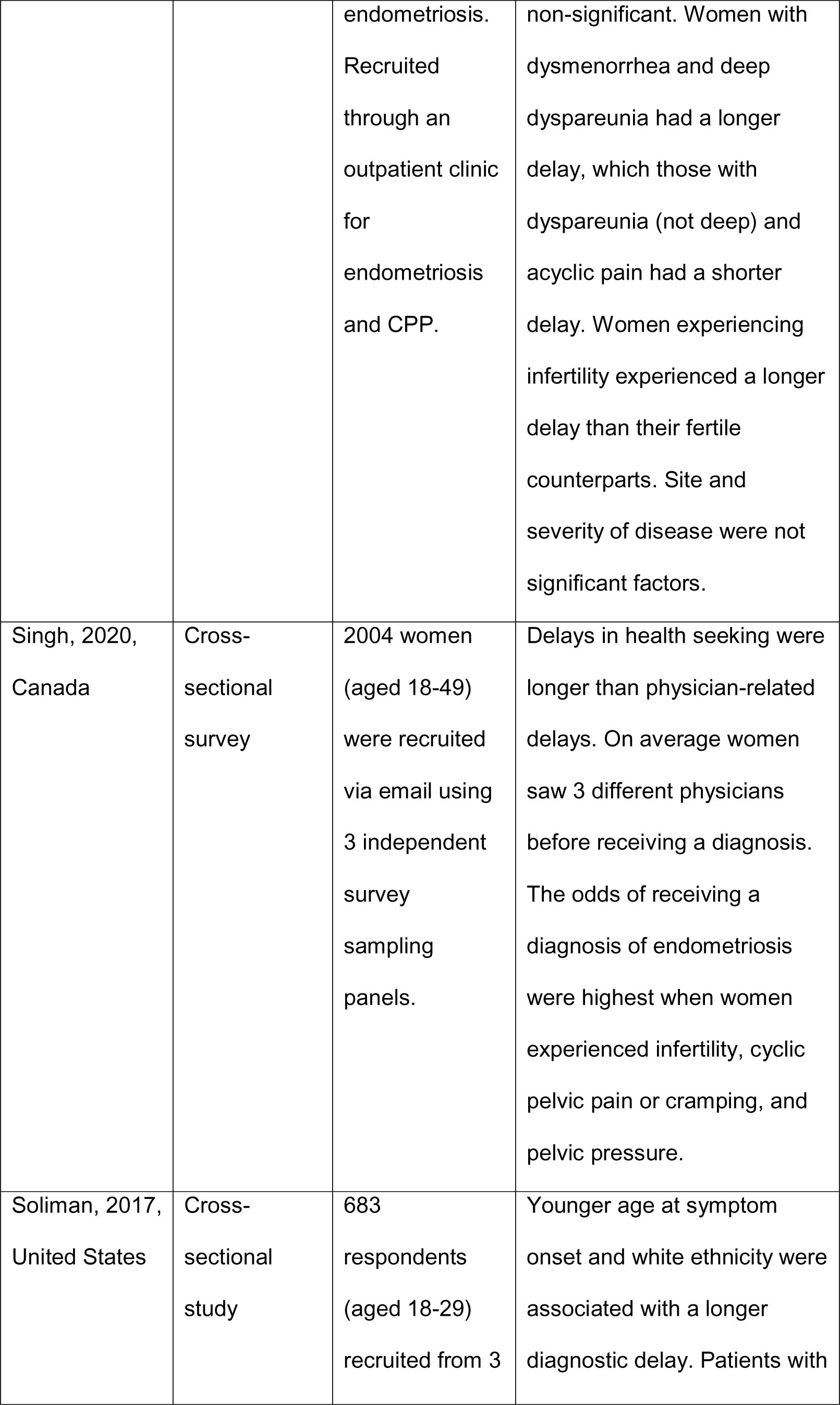
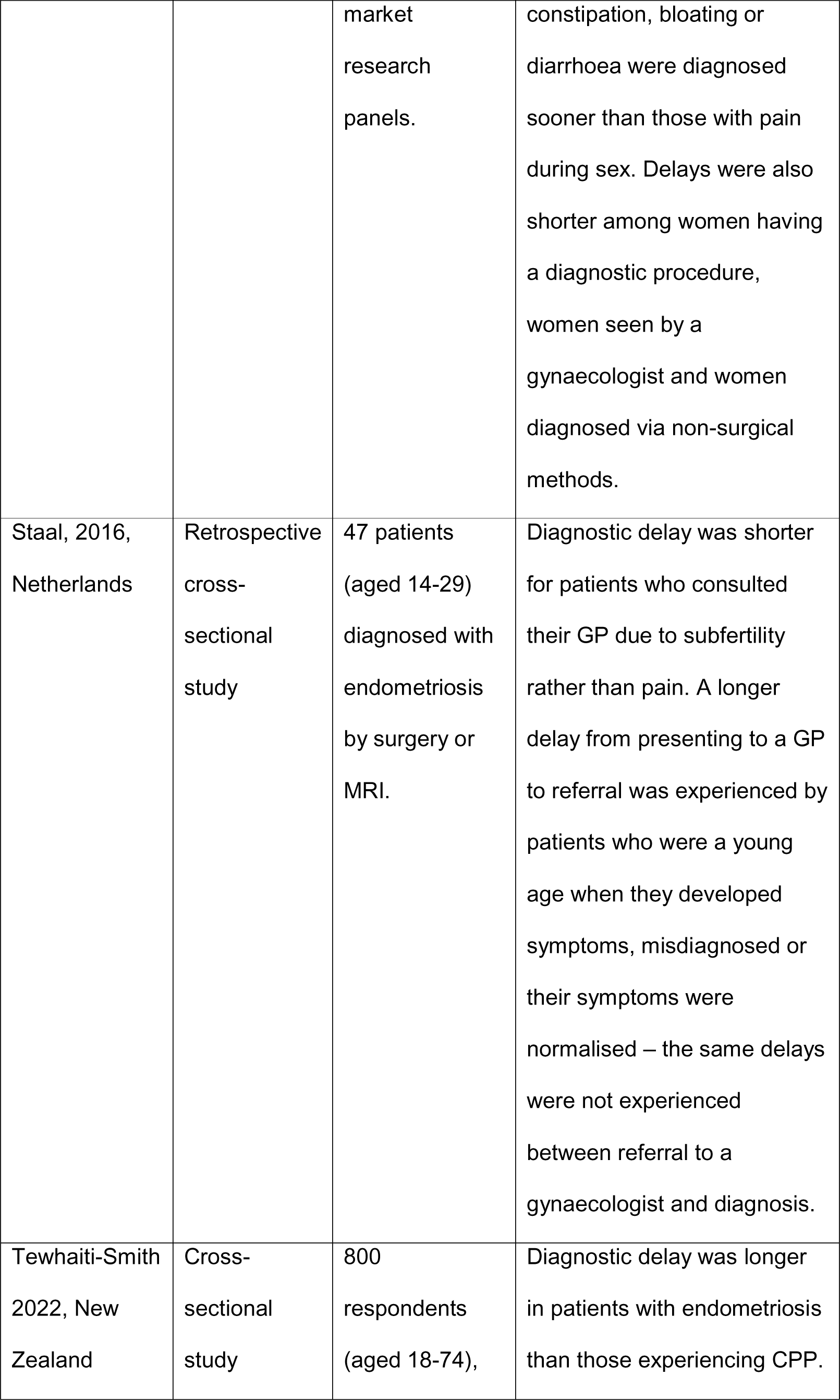
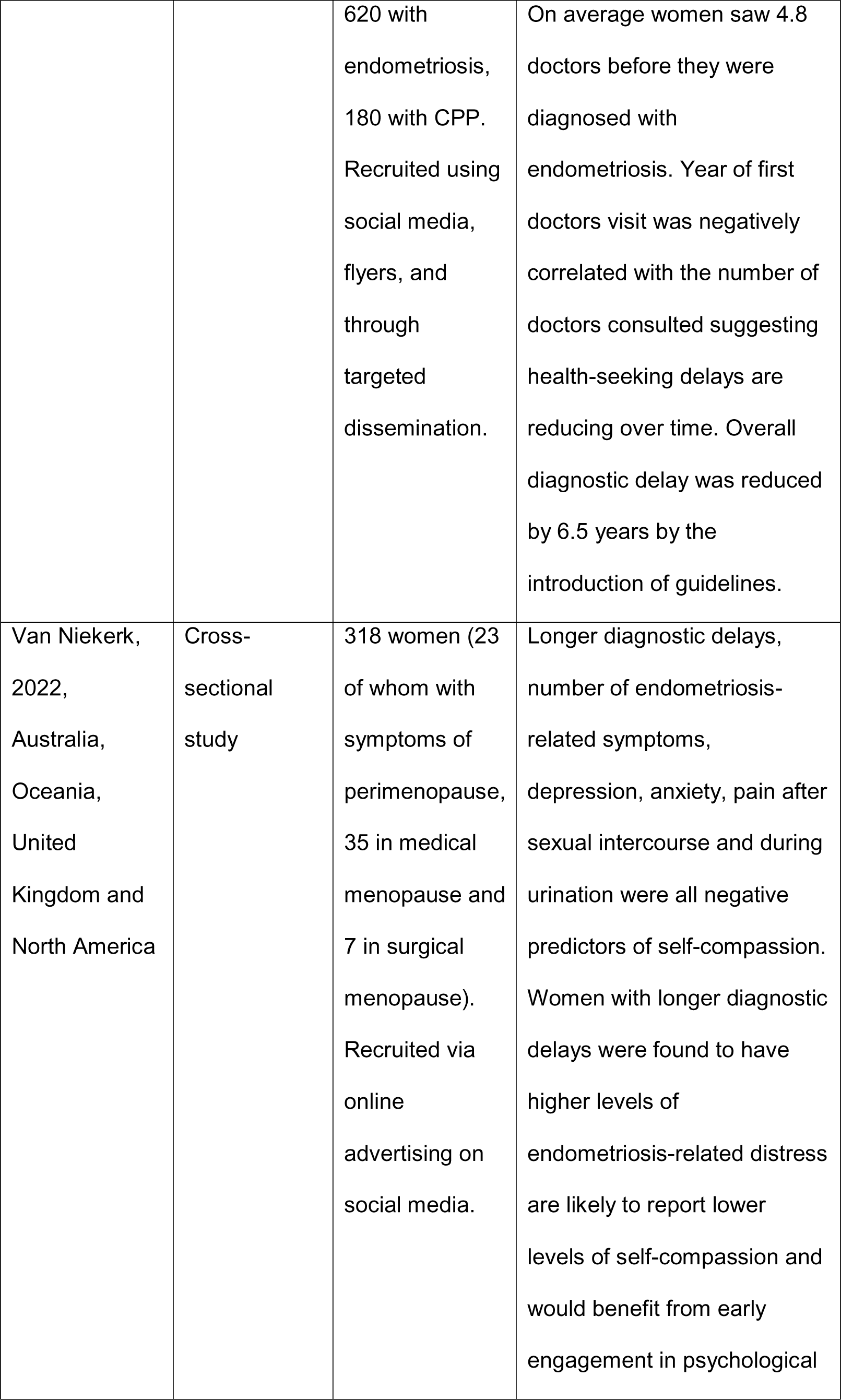
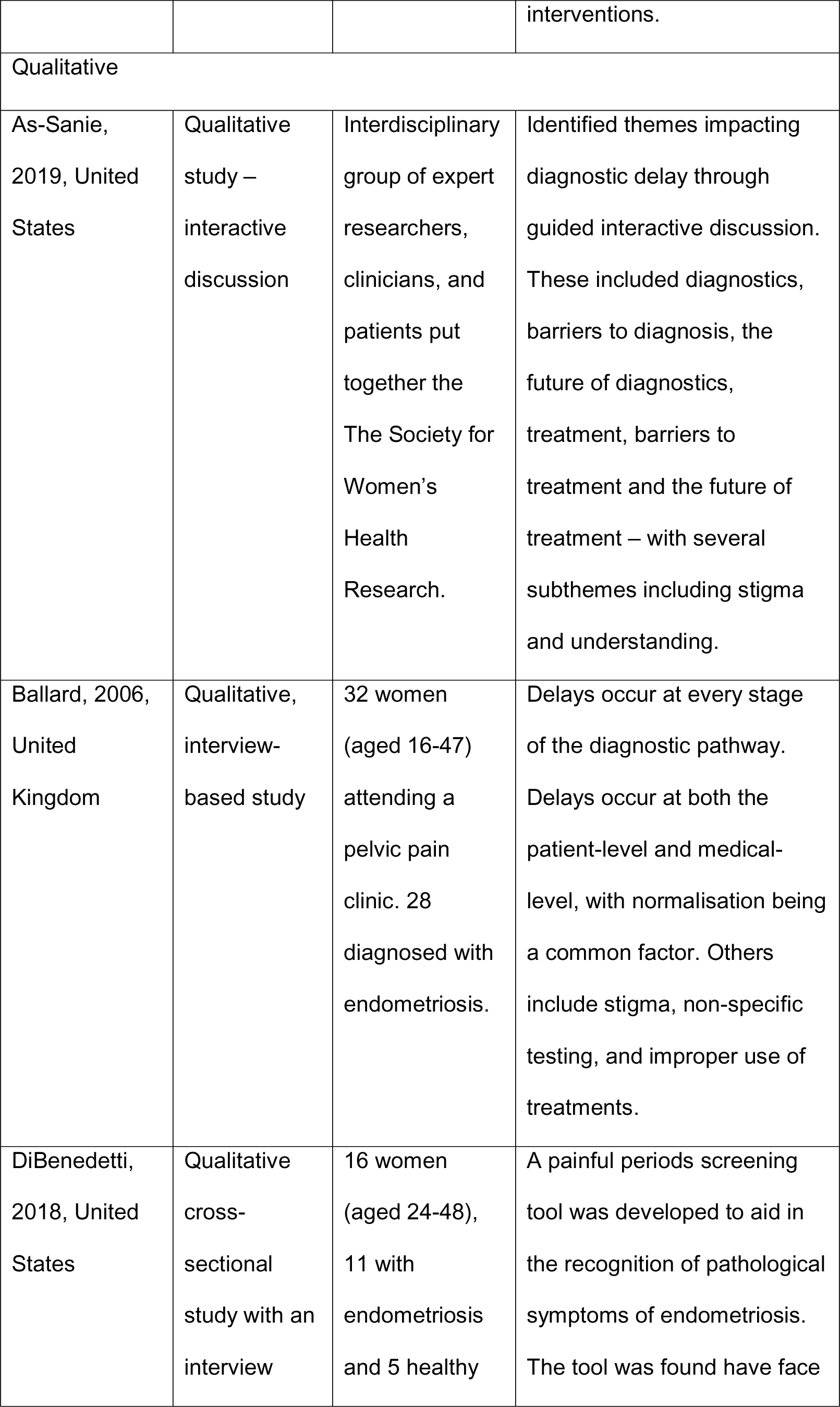
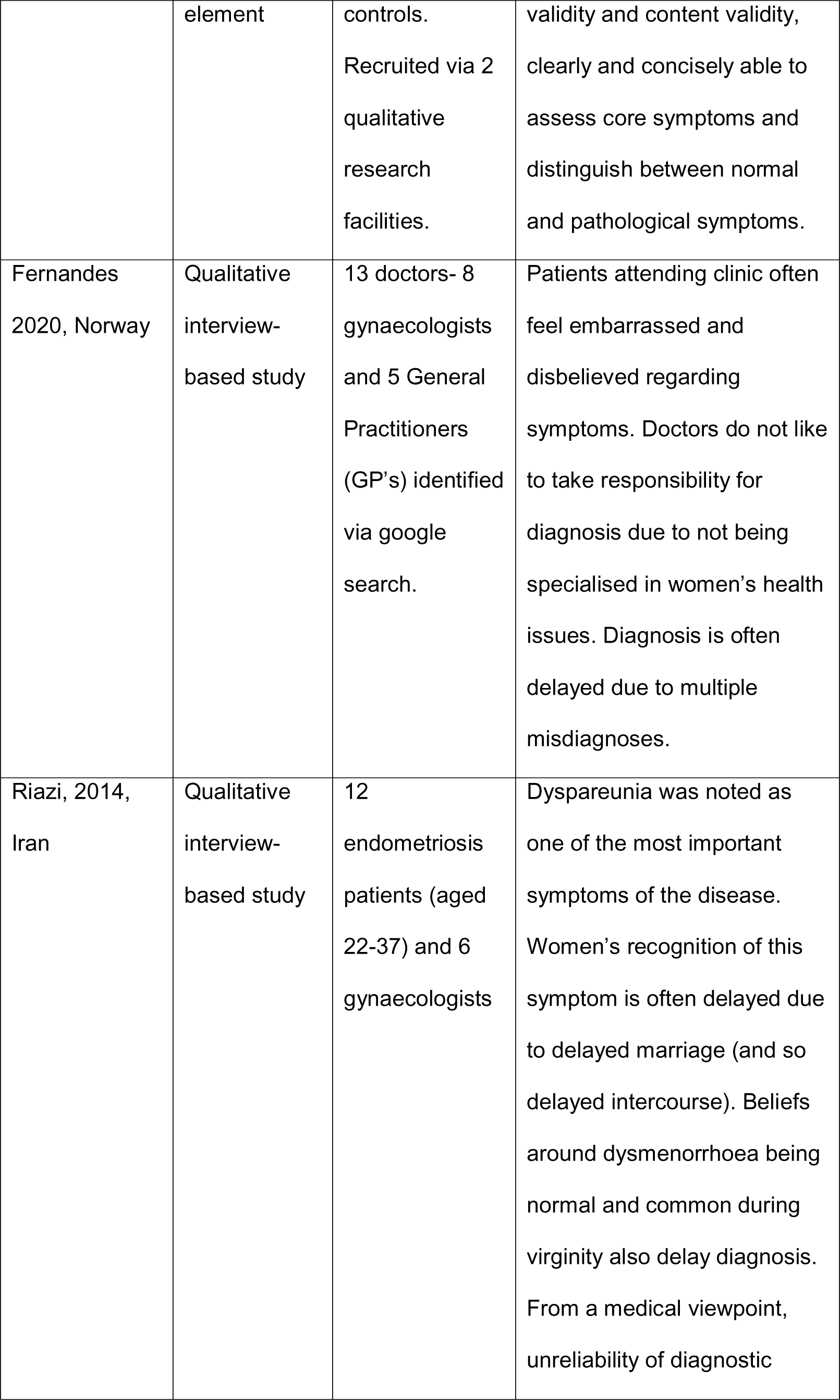
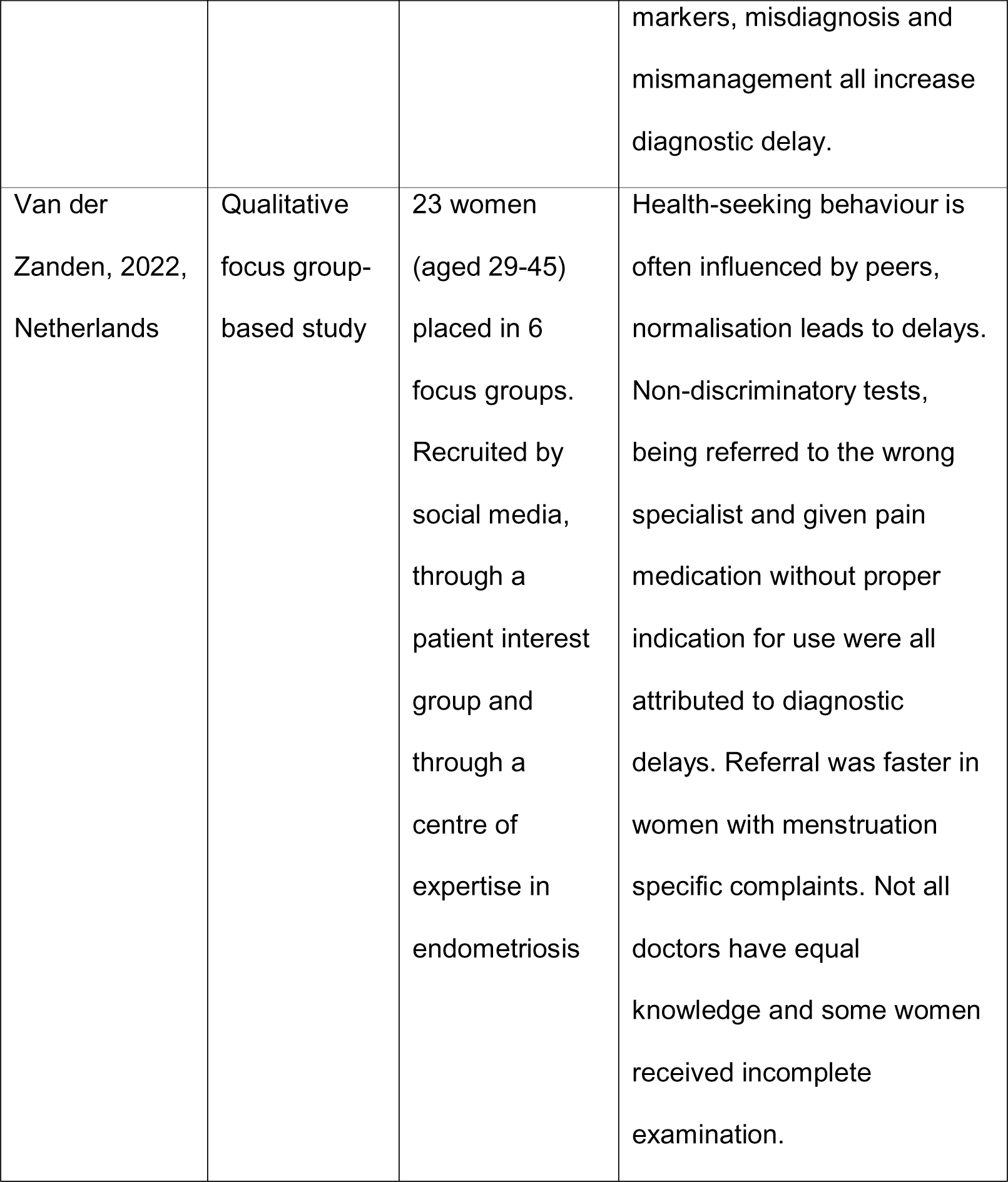
Table of included studies.

### Age of onset of endometriosis

The mean age at onset of endometriosis symptoms was 14.1 years old for adolescents (range 13-15.3 years), and 20.4 years old for adults (range 20-23.2 years). The average age at diagnosis was 16 for adolescents and 28.8 for adults (range 22-32). Average age of first GP visit was 14 for adolescents and 25.8 years for adults (range 20-32.6).

### Diagnostic delay

The definition of diagnostic delay was consistent across studies and was defined as the time between symptom onset and diagnosis. The average diagnostic delay was 6.6 years with an average of 1.5 years in Australia (27) and 11.3 years in the US (18). However, there was a wide range between the shortest and longest delay reported. The shortest delay was 0.5 years in Brazil (30), and the longest delay was 27 years in the UK (15). Though the range was wider than previously reported by other studies i.e. 3.3 - 11.7 years, the average diagnostic delay was consistent with their finding of 6.7 years (35). Some studies reported specific points at which delays occurred, these were from symptom onset to primary care consultation (15, 17–19, 21–23, 25, 26, 28, 33), referral for gynaecology consultation (15, 16, 22, 23, 33), and gynaecology referral to diagnosis (15, 16, 22, 23, 33). Mean delays through this pathway reported across the studies were 2.0 years, 2.5 years, and 2.8 years respectively. Time from primary care presentation to diagnosis was reported by some studies without mention of transition to secondary care (16, 19, 21, 25, 26). The average diagnostic delay between primary care presentation and diagnosis was 2.9 years (see figure 2).

**Figure 2:**
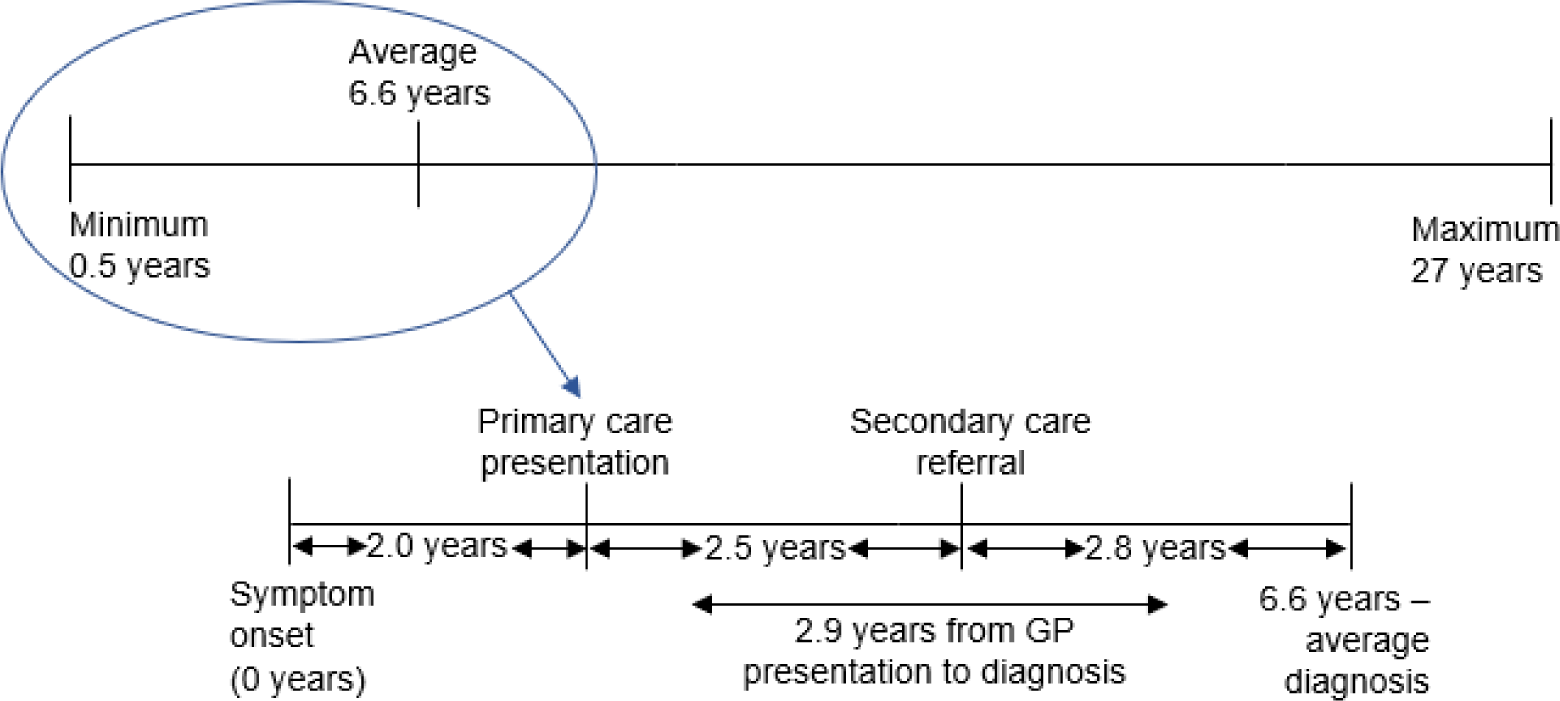
Schematic representation of global average diagnostic delay.

### Reasons for delay

Most studies focussed on the patients’ perspective, two studies focussed on the health care provider (HCP) perspective, and one included both perspectives. There were 6 main themes that emerged. A summary of these can be seen in table 2.

**Table 2:** The main themes and sub-themes relating to diagnostic delay.

|  | Main theme | Contributing factors (sub-themes) |
| --- | --- | --- |
| 1 | Access to healthcare | Physical access to care, financial barriers, stigma, embarrassment, not being aware of endometriosis, religious beliefs, and normalisation of symptoms. |
| 2 | Knowledge limitations | Poor recognition of symptoms (patients and HCP), HCP thinking endometriosis is a 'rare' disease, inability to define between normal and pathological symptoms (patients and HCP), lack of awareness and lack of training and evidence available to HCP. |
| 3 | Misdiagnosis | Differential presentation of symptoms between women, atypical symptoms, comorbidities, communication challenges between different HCP, lack of specificity of testing, lack of definitive diagnostic testing, and use of non-definitive tests. |
| 4 | Stigmatisation | Stigma, normalisation, dismissal, patient unable to properly verbalise pain and/or symptoms causing communication challenges between patient and HCP, and lack of patient assertiveness. |
| 5 | Method of diagnosis | Hesitation to refer for more invasive, definitive tests, age, HCP uncomfortable with requirement to perform physical exam (particularly adolescents), and perceived need for surgical over clinical diagnosis in some health systems. |
| 6 | Lack of guidelines | No screening tools available, inconsistency in available PROMs and guidelines, poor interdisciplinary handling of patients, and need for involvement of multiple HCP. |

Access to care differed depending on the specific health system in place. While financial barriers were more prominent for those seeking private healthcare, physical access to care was more frequently noted for those seeking public healthcare services. Only one study compared wait times between those seeking public healthcare and insurance or self-funded healthcare (35). They found that wait times for endometriosis care were significantly longer for those seeking public rather than private healthcare (8.3 years vs. 5.5 years).

Both HCPs and patients shared similar views on the reasons for diagnostic delay although they expressed the delays differently. Where HCP thought frequently presenting patients were somatising, patients stated they presented frequently because they felt unheard by HCPs. This was reflected by the number of doctors seen, which averaged 2.0 for adolescents (19) and 4.1 for adults (range 2.5-7) (19, 26, 28, 33, 35) and the number of times symptoms were discussed before diagnosis, with more than a quarter of women saying they discussed symptoms more than 20 times (34). Interestingly, none of the studies evaluated the number of consultations prior to referral, nor the effect of diagnostic delay qualitatively or quantitatively based on the type (doctor, nurse, etc.) or gender of the HCP.

The emerging themes identified increased diagnostic delay at each point along the diagnostic pathway, from symptom onset to diagnosis. This resulted in prolonging diagnosis which led to increases in both the severity of pain and the extent of disease (20, 21) (see figure 3). Both patients and HCPs appeared to demonstrate an overall lack of understanding and education about endometriosis.

**Figure 3:**
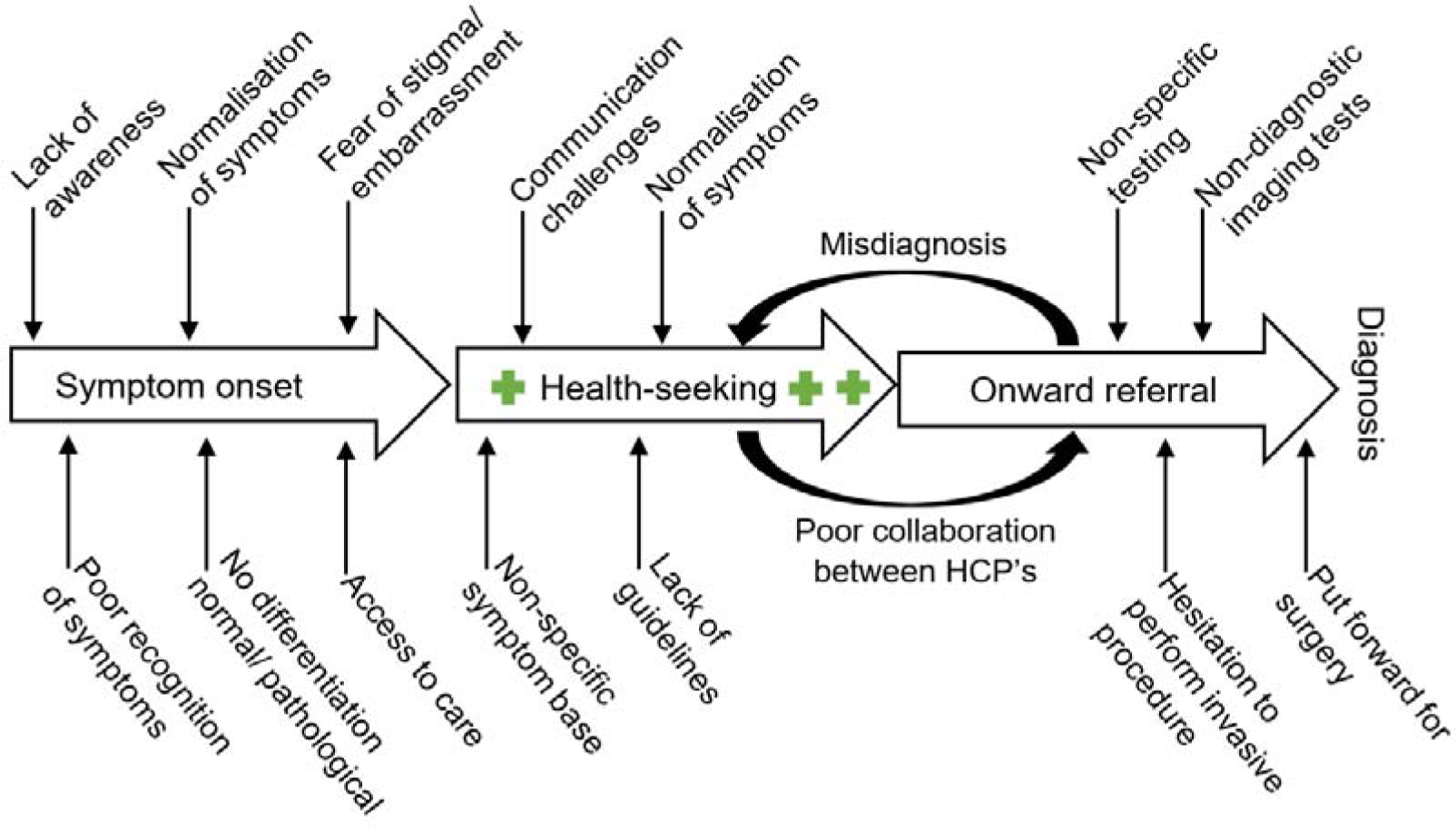
Pathways to diagnostic delay.

**Figure 4:**
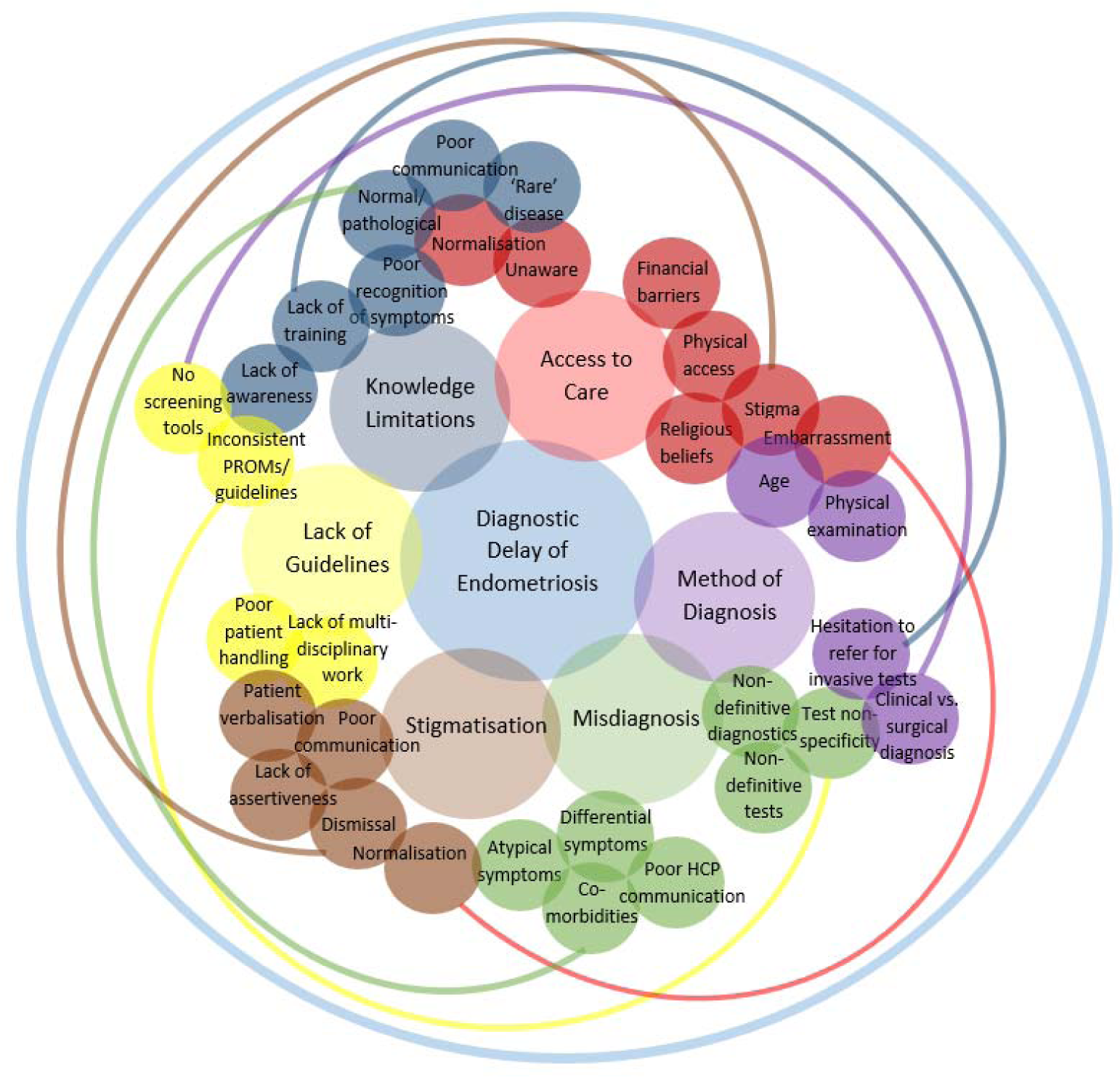
Thematic map of interactions between themes and subthemes.

### Diagnostic delay: evidence of delays being addressed

Overall, four studies reported interventions implemented to tackle diagnostic delay. Of these, two studies reported reduced time to diagnosis following the introduction of clinical guidelines (27, 28) and one study that found diagnostic delays were reduced by the introduction of specialist endometriosis centres in the US, but not in the UK (16). Only one study quantified the reduction in delay (8.4 years), while the others reported a ‘downward trend’ in diagnostic delays (16, 27, 28). Becoming a member of an endometriosis society had no effect on diagnostic delay (35).

The discrepancy in effectiveness of the introduction of specialist endometriosis centres may be due to differences in health care systems including access to care, service use, service cost, referral pathways and diagnostic guidelines.

A range of interventions to reduce diagnostic delay for endometriosis were suggested including education and awareness campaigns, collaboration, and multidisciplinary working between HCPs, promoting health-seeking behaviour for patients, the use of screening tools, increased research into endometriosis, improving access to medical records, clinical guidelines written in the native language, the use of reliable diagnostic indicators and early intervention.

The interventions suggested span the entirety of the socio-ecological framework (see figure 5). This multi-level approach to intervention allows for the introduction of all encompassing, yet targeted and effective interventions tailored according to individual factors and behaviours (13) and the wider health care system. Using this framework for diagnostic delay in endometriosis is useful to visualise the complexity involved whilst providing a range of options for intervention.

**Figure 5:**
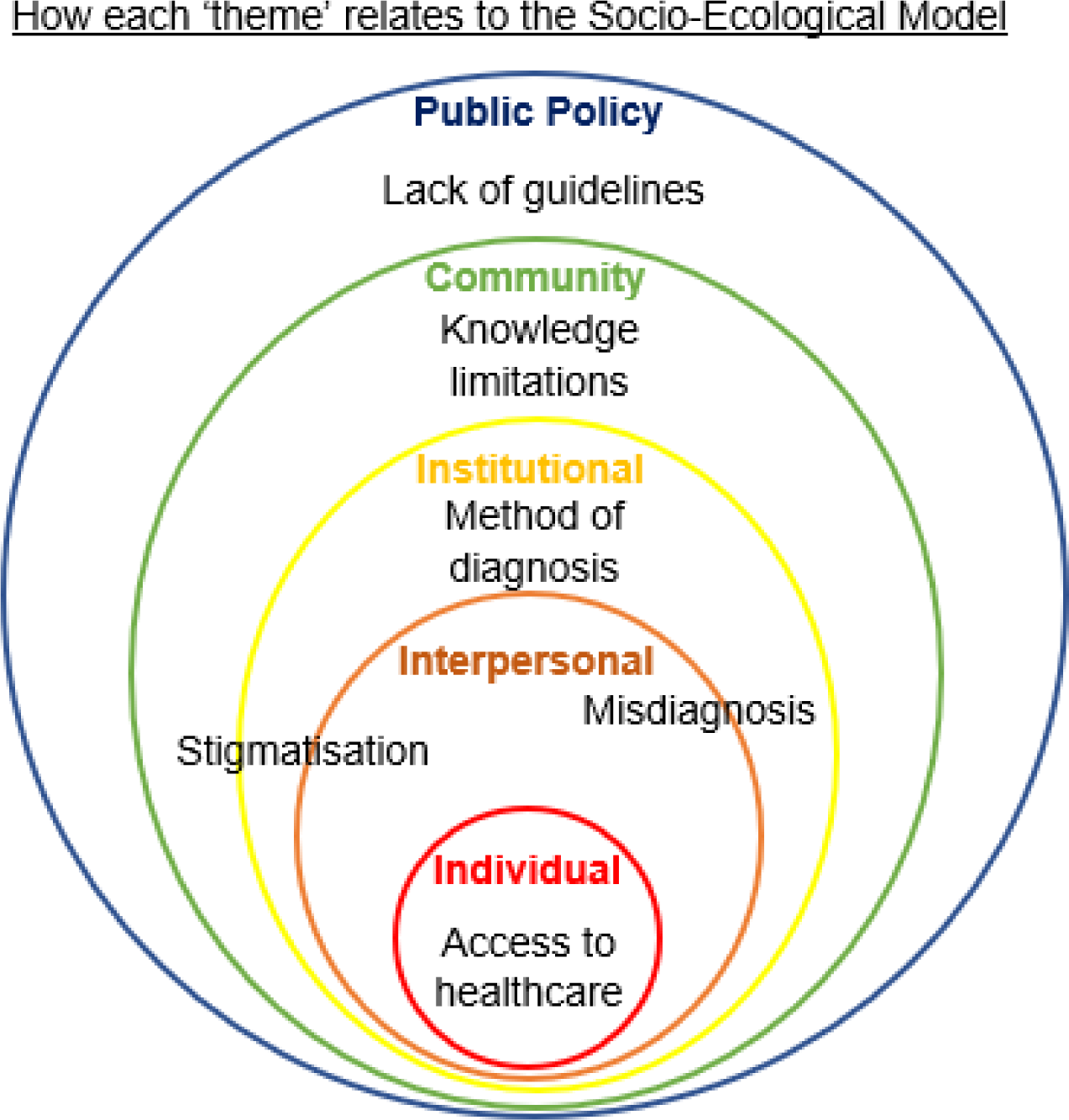
The socio-ecological model of endometriosis.

The breadth of interventions identified was aided by the diversity of participants included in the studies and was enhanced by the inclusion of views from a range of HCPs (17, 24, 32).

## Discussion

Diagnostic delay associated with endometriosis is a well-established phenomenon. Prior to our review it was not clear where in the health care system these delays occurred or why they occurred. Our review is the first attempt to synthesise and analyse this evidence. On average, the diagnostic delay for endometriosis was 6.6 years across the studies and ranged from 1.5 years to 11.3 years. Delays were identified at all stages from symptom onset to receiving a diagnosis. The longest average delay was the time from gynaecology referral to diagnosis (2.8 years), followed by primary care presentation to diagnosis (2.5 years), and finally, from symptom onset to primary care presentation (2.0 years). Only 2 studies used a CPP comparator group, while 2 used healthy controls, no other studies used a comparator or control, and none provided information on women with negative findings at laparoscopy.

We acknowledge the limitation of the scoping review methodology. The exclusion criteria meant that some papers were not included, such as those focussing on specific biomarkers. All included studies relied on patients recalling the start of their symptoms rather than tracking patients throughout their diagnostic journey or using medical records for verification which could reduce recall bias. The strength of our study was a clear focus following a pre-published protocol, including a wide range of papers from all over the world and locating the problem within the socio-ecological framework.

An area in critical need of further research is closer tracking of patients throughout their diagnostic journey. This should include the time from presentation to diagnosis, including cases where patients have met all criteria to be considered for surgery but do not have endometriosis, what their differential diagnoses are and what the differences are between women with a positive and negative laparoscopy. This may be improved by using reporting endometriosis as a differential diagnosis earlier along the diagnostic journey, and by ensuring primary and secondary care are better connected so the diagnostic journey can be properly followed. Additionally, it may be useful to have the details of the HCP available and their role e.g. primary care practitioner, gynaecologist, and their gender, age, and length of service, all of which may affect diagnostic delay.

The definition and calculation of diagnostic delay is also an area that requires urgent attention. Rather than studies describing the time from symptom onset to diagnosis, the current definition of diagnostic delay used across studies, it would be more beneficial to determine *excess delay*. This could provide regional and national estimates of the true diagnostic delay or excess delay based on regional and national average wait times for primary care appointments, referral to gynaecology, and for surgery. This measure could allow direct comparisons of care and delays between public and private provision of services for endometriosis care.

Secondly, the length of delay matters in terms of cost and severity for women and the wider health system. Accurate calculation of diagnostic delay for endometriosis may be the first step to improving guidelines, diagnostic measures, and diagnosis more broadly. Additionally, it is important to establish and address barriers to diagnosis. More investigation is needed on the effect of diagnostic delay to determine the cost-benefit of reducing diagnostic delay (37).

Though there remains much to be done, the results of this study can provide a platform for further future research to prevent the unnecessary and extended suffering resulting from diagnostic delays of endometriosis. The socio-ecological framework can be used to assess where improved policies may be effective, how widespread the effects might be and to provide a benchmark for their perceived benefit (financial and otherwise). Further research studies would benefit from utilising medical records to track the number of consultations, range of HCPs, and time elapsed from initial referral to a final diagnosis and treatment. Our review provides a starting point for others to improve our understanding of where changes need to be made to reduce the delay in diagnosis of endometriosis.

## Data Availability

All data produced in the present work are contained in the manuscript.

## Acknowledgements

We thank our employers for supporting this work. JF received support from Harrogate Borough Council, AMJ received support from the University of York and AW received support from York St John University.

